# Chronic use of a sensitized bionic hand does not remap the sense of touch

**DOI:** 10.1101/2020.05.02.20089185

**Authors:** Max Ortiz-Catalan, Enzo Mastinu, Sliman J. Bensmaia

## Abstract

Electrical stimulation of tactile nerve fibers that innervated an amputated hand results in vivid sensations experienced at a specific location on the phantom hand, a phenomenon that can be leveraged to convey tactile feedback through bionic hands. Ideally, electrically evoked tactile sensations would be experienced on the appropriate part of the hand: Touch with the bionic index fingertip, for example, would elicit a sensation experienced on the index fingertip. However, the perceived locations of sensations are determined by the idiosyncratic position of the stimulating electrode relative to the nerve fascicles and thus difficult to predict or control. This problem could be circumvented if perceived sensations shifted over time so that they became consistent with the position of the sensor that triggers them. We show that, after long term use of a neuromusculoskeletal prosthesis that featured a mismatch between the sensor location and the resulting tactile experience, the perceived location of the touch did not change.

## Main text

Manual interactions with objects give rise to a barrage of neural signals from the skin about the objects themselves – their size, shape, and texture – and about our interactions with them – contact timing, force, and location (Johansson and Flanagan, 2009). Without these tactile signals, dexterous manipulation would be severely impaired, as evidenced by the deficits resulting from digital anesthesia or deafferentation (Johansson *et al*., 1992). The importance of tactile feedback in manual behavior has spurred the development of strategies to convey tactile signals in bionic hands. One promising approach to sensory restoration is to establish an electrical interface with the residual nerve through chronically implanted electrodes, as microstimulation of the nerve evokes vivid sensations experienced on the phantom hand (Clippinger *et al*., 1974; Ortiz-Catalan *et al*., 2014; Tan *et al*., 2014; George *et al*., 2019; Petrini *et al*., 2019).

In principle, the more naturalistic are these artificially induced neural signals, the more intuitive will the resulting sensations be (Saal and Bensmaia, 2015; Valle *et al*., 2018; George *et al*., 2019). The most straightforward application of this principle of biomimicry is somatotopic mapping: As stimulation through a given electrode evokes a percept that is localized to a specific patch of skin, connecting a sensor on the corresponding part of the bionic hand to that electrode is likely to convey intuitive information about contact location (Saal and Bensmaia, 2015). For instance, if stimulation through an electrode gives rise to a sensation on the index fingertip, it stands to reason to connect the index fingertip sensor to that electrode: Anytime the bionic index fingertip touches an object, the subject will experience a sensation on their fingertip and will thus know where contact was initiated without having to think about it (Dhillon and Horch, 2005).

The problem with the somatotopic mapping strategy is that, in practice, the projection field associated with each electrode – the region of the phantom on which the sensation is experienced when current is delivered through that electrode – is idiosyncratically determined by the location of the electrode on or in the nerve, and cannot be prearranged by the implanting surgeon. As a result, a given electrode array may not impinge on some hand regions – one or more fingertips, e.g. – where most contact with objects occurs (Christel *et al*., 1998).

If one cannot control the location of the projection fields, one might hope to relocate them after implantation. Indeed, when the limb region of somatosensory cortex is deafferented through amputation, this deafferented cortex can be activated via touch applied to other body regions (Pons *et al*., 1991) and amputation of a digit leads to an increase in the neural territory that can be activated through tactile stimulation of adjacent digits (Merzenich *et al*., 1984). These findings have been interpreted as evidence that body maps may be malleable. Moreover, the repeated pairing of a tactile experience with a visual touch applied to an extracorporeal object leads to the fusing of the two sensory experiences (Botvinick and Cohen, 1998). One might thus hope that the visual experience of touching one part of the bionic hand – where a touch sensor is located – paired with a timely tactile sensation to another part of the phantom hand – the projected field of an electrode – will lead to a shift in the perceived location of the sensation, driven by a reorganization of the body map in the brain.

To test this hypothesis, three unilateral transhumeral amputees were instrumented with a neuromusculoskeletal prosthetic arm and hand (Ortiz-Catalan *et al*., 2014, 2020) (Figure 1A). The hand was controlled via electromyographic signals measured using electrodes implanted on the muscles (Ortiz-Catalan *et al*., 2012). Moreover, tactile feedback was conveyed by electrically stimulating the median or ulnar nerves (Mastinu *et al*., 2017). Activation of a sensor located on the prosthetic thumb drove electrical stimulation through one electrode contact – dubbed here the “feedback contact” – implanted around the ulnar (Participant 1) and median (Participants 2 and 3) nerves. Participants lived with this closed-loop myoelectrically controlled bionic hand and used it to performed activities of daily living for up to three years.

Participants wore the prosthesis every day while awake, except when showering or swimming, based on verbal reports and onboard usage tracking (mean daily usage hours: 18.4, 15.4, and, 13.1 hours for P1, P2, and P3, respectively). The hand was actuated throughout the day as well, as evidenced by tens of minutes of use for each participant (Figure 1B), implying more than 100 grasping actions per day (assuming each grasping movement lasts an average of a few seconds).

**Figure 1.**
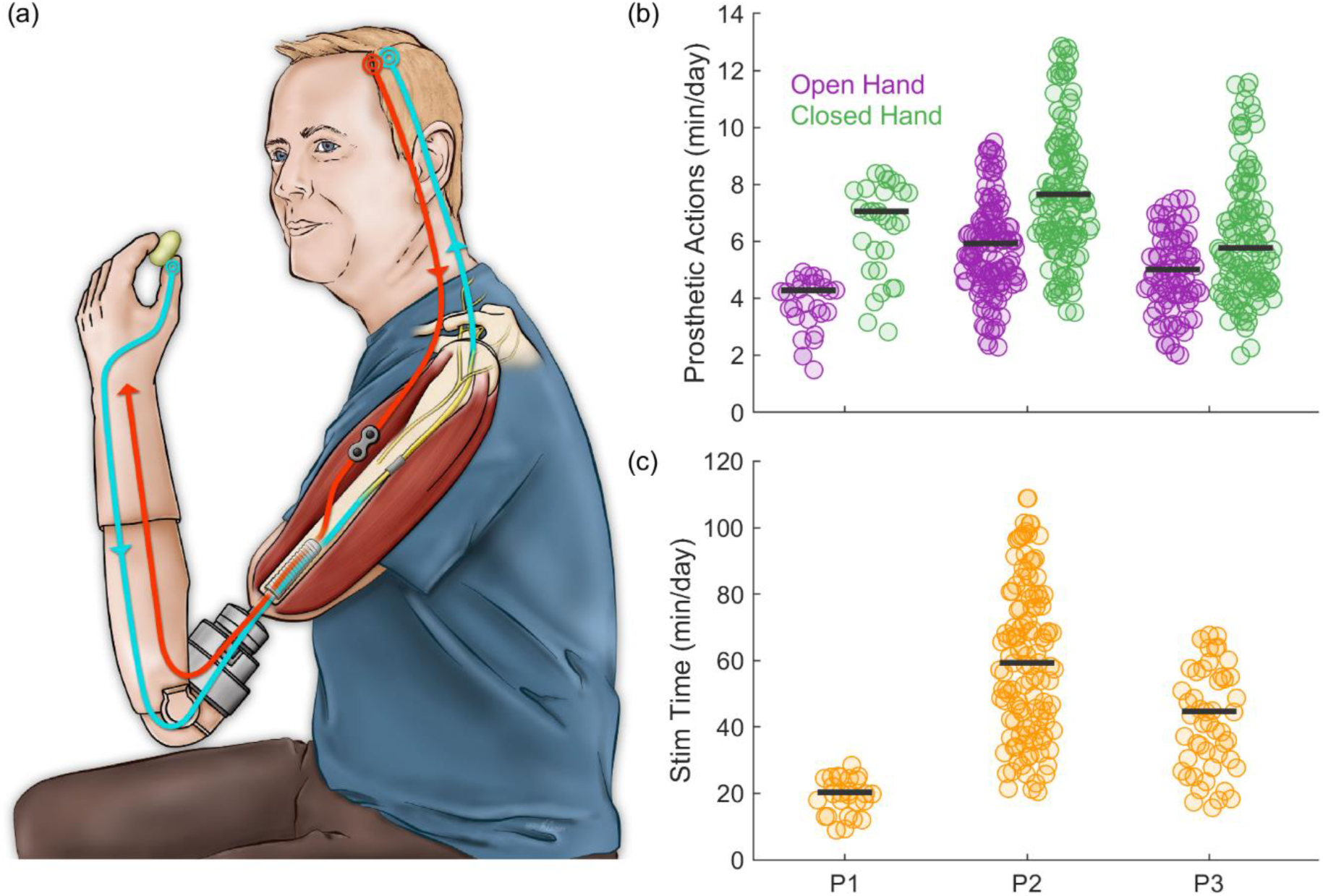
Neuromusculoskeletal prosthesis used in daily life. A| Participant wearing a neuromusculoskeletal prosthesis connected to his skeleton, nerves, and muscles. Implanted electrodes on muscles and nerves are used for control (red) and sensory feedback (blue), respectively. The interface between internal and external components of the bionic hand is through an osseointegrated implant into the bone. B| Cumulative time of prosthetic actuation. C| Cumulative time per day of neurostimulation for each of the three participants. The prosthesis was worn all the time the participants were awake during the day.

Contact with the prosthetic thumb resulted in electrical stimulation of the nerve for up to 5 seconds at a time (the duration was capped for safety reasons). The frequency of the electrical stimulation was graded according to the sensor output to modulate the perceived magnitude (Ortiz-Catalan *et al*., 2020.; Dhillon and Horch, 2005; Tan *et al*., 2014; Graczyk *et al*., 2016), thereby conveying information about applied pressure. All three participants experienced extensive stimulation each day (mean daily stimulation duration: 20.3, 59.3, and 44.7 minutes for P1, P2, and P3, respectively, see Figure 1C).

The prosthetic hand allowed for superior grasping force precision and reliability when compared with conventional surface electrode control (Mastinu *et al*., 2019). Moreover, the sensory feedback proved beneficial for restoring grasping coordination and assisting corrective actions when grasping under uncertainty, for example when the weight of the object changed unexpectedly (Mastinu *et al*., n.d.). Additionally, long-term home-use of the tactile sensory feedback led to increased sensitivity to changes in electrical stimulation, as evidenced by improved pulse frequency discrimination (Ortiz-Catalan *et al*., 2020.). Furthermore, participants reported greater confidence in their prosthesis control as well as improved self-image and self-esteem, leading to better social relationships and increased participation in a wider range of activities. Participants also expressed increased embodiment of the bionic limb, claiming that it is “part of my body,” “it is my arm now,” or “I don’t carry it; it is me” (Ortiz-Catalan *et al*., 2020).

For at least one year prior to enabling electrical stimulation of the nerve, participants used their bionic hand without sensory feedback (Ortiz-Catalan *et al*., 2014). During this period, we tracked the location of the projected field of the different contacts on the cuff electrode. To this end, we periodically delivered microstimulation pulse trains through one of several contacts (3, 6, and 5 for P1, P2, and P3, respectively) – interleaved in random order – and interrogated the subject as to where the sensation was experienced. Results from the full mapping are reported elsewhere (Ortiz-Catalan *et al*., 2020; Ackerley *et al*., 2018). Here, we present results for the feedback contact, which was paired with the sensor.

Feedback contacts had projected fields located on the hypothenar (P1), proximal fingerpad of the thumb (P2), and the distal fingerpad of the middle finger (P3) (Figure 2A). The location of these projected fields remained consistent over repeated testing during the year preceding the pairing with the sensor (blue hues, Figure 2A,B). More importantly, and perhaps surprisingly, the location of the projected field did not change after pairing with the sensor (green hues, Figure 2A, B). That is, over the period of over one year, every time the participants’ prosthetic thumb contacted an object, they experienced a tactile sensation somewhere else on the hand and the location of that tactile sensation did not change. Periodic testing of the location of the projective field showed that it moved only slightly – typically a millimeter or less – from test to test (typically separated by weeks or months) (Figure 2B). Furthermore, the direction in which the projected field moved was random, as evidenced by vector strengths that were not significantly different from those expected if the direction of movement was uniformly distributed (Figure 2C).

**Figure 2.**
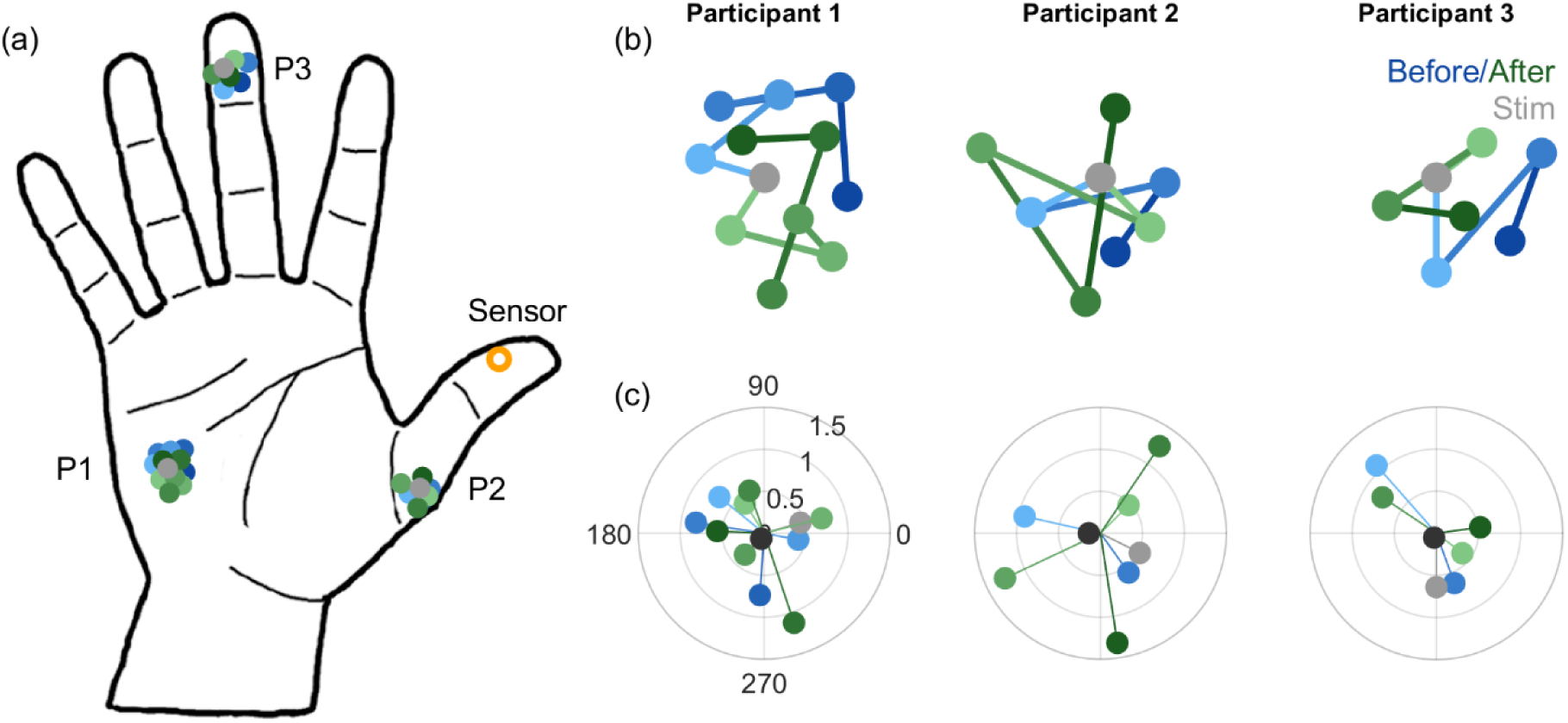
A| Location of the projection fields of the feedback electrodes over the course of the study. Gray dots represent the location of the projection fields measured on the session when sensory stimulation was activated for home use. Shades of blue and green illustrate the locations before and after this pairing, respectively. Time periods varied across participants: Participant 1 (P1), 27 months before and 28 months after stimulation; Participant 2 (P2), 10 months then 12 months; and Participant 3 (P3), 6 months then 12 months. B| Sequence of projected fields zoomed in for each participant. C| Angle and extent of the displacement of the projected field in consecutive measurements. One would expect the angle to be consistent if it was moving systematically toward the sensor, but angles were random (vector strength was not significantly different from what would be observed in the direction and extent of movement was random from measurement to measurement).

The location of the projection field of the feedback contact was thus remarkably stable, despite the chronic mismatch between the visual experience of contact location and its tactile counterpart. This fixedness is especially surprising given that the prosthesis was used on a daily basis and the sensory feedback was behaviorally relevant (Recanzone *et al*., 1993). The possibility remains that the visuo-tactile mismatch was not salient enough to promote plasticity. Indeed, the participants may not have looked at their bionic hand frequently enough to experience the visuo-tactile mismatch. Or perhaps contact timing during typical object interactions is consistent enough across bionic fingers that this mismatch was obscured. However two of the three projection fields were not on fingertips, so this is unlikely. Given these caveats, we cannot exclude the possibility that another approach to remap projection fields, for example by repeatedly pairing the electrical stimulus with a visual cue at the desired location of the projection field (Rognini *et al*., 2018), would lead to a remapping of the sensory experience. Furthermore, the projection field and the sensor location can become more tightly aligned through chronic home use of a prosthesis if these are somatotopically mapped to begin with (Cuberovic *et al*., n.d.; Schofield *et al*., 2020).

Nonetheless, our results suggest that the visuo-tactile mismatch does not resolve itself when participants perform activities of daily living with the bionic hand, even over an extended period. This finding is consistent with a view that sensory maps are highly stable in adulthood (Makin and Bensmaia, 2017) and cannot be meaningfully modified, even with prolonged exposure.

## Materials and Methods

### Participants and the neuromusculoskeletal arm prosthesis

Three participants with transhumeral amputation implanted with a neuromusculoskeletal arm prosthesis participated in the study. Details on the participants medical background is provided in reference (Ortiz-Catalan *et al*., 2020). Participant 1 (P1) was implanted in 2013 (Ortiz-Catalan *et al*., 2014), Participants 2 and 3 (P2 and P3) in 2017. P2 and P3 underwent a targeted muscle reinnervation (TMR)(Kuiken *et al*., 2009) surgical procedure aimed at providing intuitive myoelectric signals for hand opening and closing. The neuromusculoskeletal interface (e-OPRA, Integrum AB, Sweden) consists of 1) an osseointegrated percutaneous titanium implant for direct skeletal attachment of the artificial limb, 2) feedthrough connectors embedded in the osseointegrated implant to allow the artificial limb to communicate with implanted electrodes, and 3) implanted electrodes in nerves and muscles with up to 16 electrode contacts (Ortiz-Catalan *et al*., 2020). Epimysial electrodes were sutured on both naturally innervated and surgically reinnervated muscles, and spiral cuff electrodes were wrapped around the ulnar nerve for P1, and the ulnar and median nerves for P2 and P3 (Ortiz-Catalan *et al*., 2012). A custom-designed embedded electronic system placed at the interface between the neuromusculoskeletal interface and the prosthesis was used for signal processing, control, and neurostimulation (Mastinu *et al*., 2017). The study was approved by the Swedish regional ethical committee in Gothenburg (Dnr: 769-12) and all participants provided written informed consent.

### Prosthetic setup and control

The prosthetic setup for all participants consisted of a myoelectric hand (SensorHand, Ottobock, Germany), elbow (ErgoArm, Ottobock, Germany) and the artificial limb controller (ALC), a custom-designed embedded system for closed-loop prosthetic control that serves the dual purpose of recording EMG to control prosthesis movement and providing sensory feedback via neural stimulation (Mastinu *et al*., 2017). The prosthesis was self-contained and did not require external batteries, processing, or stimulation equipment. Myoelectric signals from the epimysial electrodes were sampled at 500 Hz, high-pass filtered at 20 Hz, low-pass filtered at 250 Hz, and notch-filtered at 50 Hz. The prosthetic hand was commanded using direct control (also known as one-for-one control), where the mean absolute value of an EMG channel (over a 100-ms time window) was proportionally mapped to the actuation speed. The thresholds for direct control were customized for each participant to provide optimal control of the terminal device.

### Sensory feedback for home-use

Participants were provided with tactile sensory feedback for home-use in January 2017 (P1) and September 2018 (P2 and P3). Electrical stimulation of the residual nerves via cuff electrodes depended to the output of three sensors located on the prosthetic thumb. The average readout of the force sensors was linearly mapped to the pulse frequency within the range from 5 Hz to 30 Hz (Günter *et al*., 2019). Stimulation stopped when the sensors were no longer in contact with an object or after 5 seconds, whichever happened first.

Stimulation pulses were cathodic-first, rectangular, bipolar (50 μs inter-pulse delay), asymmetric (10:1), charge-balanced, and current-controlled. Only one contact of the cuff electrode per participant was used for home-use stimulation, prioritizing ones that required the least charge to elicit perception, that is, the ones yielding the lowest detection threshold. Perceptual threshold was measured by delivering single pulses at different amplitudes and widths and having subjects report whether or not they felt the stimulus. Charge was gradually increased until the subject reported a tactile percept. This procedure was repeated on all the electrodes and the pulse width yielding the lowest charge threshold was identified for each electrode

The projected field – the location at which a tactile percept was experienced – was reported by the participant by marking it on an image as that shown in Figure 2A in the main text. For these measurements, threshold level stimulation was delivered, which resulted in highly localized tactile percepts, reported by the participants to feel like “being touched with the tip of a pen.” Electrical stimulation was never reported as painful.

### Characterizing the progression of projected fields

To characterize the progression of the projected fields over time, we first plotted their trajectory in two dimensions (Figure 2B). We then produced a polar plot of the displacement direction and extent between each measurement (Figure 2C). We could then assess whether the projected fields tended to move in any one direction. To this end, we computed the vector strength (Mardia, 1975), given by:

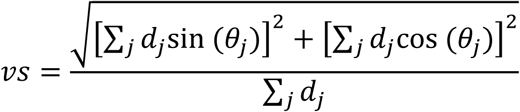

where *d_j_* is the distance over which and *θ_j_* is the direction in which the projection field moved from one measurement to the next. We then characterized using a Monte Carlo simulation the distribution of vector strengths that would be obtained if the direction and extent was randomized from step to step (by sampling them from a uniform distribution from 0 to 2π), matching the number of steps. Repeating this sampling 100,000 times, we computed the proportion of times the measured vector strength was larger than what would expected by chance, the equivalent of a p-value for each measured vector strength.

## Data Availability

The data presented in this manuscript can be made available by contacting the corresponding author with a reasanoble request.

## Acknowledgments

We would like to thank Kenzie Green for the illustration and Charles Greenspon for figure preparation and data analysis.

